# Shared and unique deficits of social functioning in multiple affected families with major psychiatric illness

**DOI:** 10.1101/2023.05.25.23290270

**Authors:** Moorthy Muthukumaran, Sowmya Selvaraj, Srinivas Balachander, Ravi Kumar Nadella, Vanteemar S. Sreeraj, Sreenivasulu Mullappagari, Shruthi Narayan, Pramod Kumar, Anand Jose Kannampuzha, Alen Chandy Alexander, Pavithra Dayalamurthy, Mahashweta Bhattacharya, Mino Susan Joseph, Sweta Sheth, Joan C Puzhakkal, Navya Spurthi Thatikonda, Dhruva Ithal, Biju Viswanath, Sydney Moirangthem, Ganesan Venkatasubramanian, John P John, Jagadisha Thirthalli, YC Janardhan Reddy, Vivek Benegal, Mathew Varghese, Sanjeev Jain, ADBS Consortium

## Abstract

**Background:** Mental illnesses often cluster in families, and their impact on affected and unaffected members within families need to be understood from a social perspective.

**Methods:** Data was derived from 202 families with multiple affected individuals, identified under Accelerator program for Discovery in Brain disorders using Stem cells (ADBS) study. Affected individuals (N=259) had a diagnosis of schizophrenia, bipolar disorder, obsessive-compulsive disorder or substance use disorder. For comparison, we used the unaffected siblings from the same families (N=229), and a matched random subset of healthy control (HC) data (N=229) from the National Mental Health Survey (NMHS). We compared educational attainment, functional marital status and occupational status between the groups.

**Results:** The groups were matched across age, gender and place of residence. The highest educational attainment was significantly different between the groups. The affected (9.9 years) and unaffected siblings (10.4 years) had poor educational attainment compared to HC (11.6 years), (F=8.97, p<0.001). Similarly, the affected (43%) and unaffected siblings (33%) remained more often single, in contrast to HC (23%), (χ2=40.98.17, p<0.001). However, employment rates were significantly higher in the unaffected siblings, especially female siblings. Females had overall lesser educational attainment, were largely married, and were majority homemakers across the three groups when compared to males.

**Discussion:** Our study findings reveal the socio-demographic characteristics of affected and unaffected siblings from multiple affected families with major psychiatric illness in India. Affected and unaffected siblings had lower educational attainment and rates of marriage when compared to HC. The unaffected siblings were more likely to be in employment compared to HC. Whether the poor educational attainment and lower marriage rates in unaffected siblings is a biological marker of shared endophenotype or the effect of social burden of having an affected family member requires further systematic evaluation.

## INTRODUCTION

Severe mental illnesses (schizophrenia, bipolar disorders, obsessive-compulsive disorder, and substance use disorders) are associated with impairment and disability in work, family and social life; may pose a significant burden on the family members. These illnesses may represent an accumulation of risk factors, which siblings may share. Thus, many attributes such as educational performance, social skills, etc., may also be a shared environmental and genetic liability .

In India, families are the mainstay of caregiving for persons with mental illness (Shankar & Rao, 2005). Co-habiting with a family member suffering from mental illness may affect various psychosocial factors, such as their functioning capacity, work conditions, emotional distress and change in family dynamics, generally leading to adverse outcomes (Rhee & Rosenheck, 2019). Mental illnesses tend to run in families due to inherited genetic and shared environmental risk factors (Antypa & Serretti, 2014). The risk of first-degree relatives developing mental illness is approximately 10-20 times higher than that of the general population (Kessler et al., 1994; Smith, Greenberg, Sciortino, Sandoval, & Lukens, 2016). The burden of care may be even higher in “multiplex families” where more than one member is affected by severe mental illnesses (Shankar & Rao, 2005). Siblings may share genetic traits, and social and environmental adversities, in their life-long relationship (Smith et al., 2016). They may also be ‘at-risk’ for general functional impairments, even when not having a diagnosable syndrome, apart from being prone to decompensate during stressful conditions (Shivers & Textoris, 2021). Additionally, these siblings are faced with the challenge of coping with increased familial burden from childhood (Marsh & Dickens, 1997; Lukens, Thorning, & Lohrer, 2004). This challenge may interfere with the sibling’s social life or require the sibling to take on additional caregiving responsibilities to help their family (Marsh & Dickens, 1997) (Lukens et al., 2004). Having a sick relative can be stigmatizing which impacts other aspects of social functioning like marriage and employment.

There is thus a need to understand the social and occupational attainment of siblings who have “multiple ill members within their family.” These patterns across generations may be confounded by secular trends, especially in developing countries where education and social facilities may differ across generations (Azam & Bhatt, 2015; Maitra & Sharma, 2009). Hence, assessment of social & occupational functioning in siblings, who are likely to have had similar opportunities, and contrasting this with individuals who do not have family affected with mental illness, can help us identify if there are any differences.

## MATERIALS AND METHODS

This is a cross-sectional comparative study from the data obtained in the larger ongoing longitudinal study, the Accelerator program for Discovery in Brain disorders using Stem cells (ADBS) project at National Institute of Mental Health and Neurosciences (NIMHANS) in collaboration with National Centre for Biological Sciences (NCBS) and the Institute for Stem Cell Science and Regenerative Medicine (InStem) (Viswanath et al., 2018; Someshwar & Holla, 2020; Sreeraj et al., 2021). Individuals with no first-degree relatives suffering from psychiatric illness from southern India, as evaluated during the National Mental Health Survey (NMHS), were the comparison group (Pradeep BS et al, 2018). The studies were approved by the NIMHANS ethics committee and written informed consent was obtained from all participants.

### Participants

#### Affected individuals and their unaffected siblings from the ADBS cohort

The ADBS study includes families in whom multiple members (at least two affected first-degree relatives in the same family) are diagnosed to have any of the major psychiatric disorders [schizophrenia, bipolar disorder, obsessive-compulsive disorder (OCD), and substance use disorder] (Viswanath et al., 2018). Recruitment of this study initiated in 2016, and at the time of this study, 2256 individuals from 472 multiplex families have been recruited. Details regarding the inclusion/exclusion criteria and the diagnosis of the index probands in the study are provided in an earlier published report (Sreeraj et al, 2021). We used a subset of this cohort, with families in whom data of sibling pairs with at least one affected and one unaffected sibling were available. All included individuals were assessed by trained mental health professionals, and were clinically examined by at least two psychiatrists.

#### National Mental Health Survey of India (NMHS) Dataset

The National Mental Health Survey of India sampled a total of 34,802 participants across the country. From this, we derived a subset of healthy participants (without a psychiatric illness) from the southern Indian states - Kerala and Tamil Nadu (N=3342), as most subjects under ADBS are from these regions. To obtain a subset of age and gender-matched controls from the NMHS sample, we used propensity score matching (detailed below).

#### Outcome measures

We compared the highest attained education, functional marital status and occupational status among three groups. The participants were grouped for the functional marital status as “single” if they were unmarried; if the couple were legally divorced or married but not living together due to estrangement, indicating they were unable to sustain a marital relationship as “divorced or separated.” Those who remained married or widowed were considered as “married.” The individuals were classified as “employed” if they were employed in a government or private sector fetching a fixed income, or self-employed without any fixed monthly income; as “student” if the person was pursuing any course and not receiving any wages; as “homemaker” if the person is managing the household affairs without any intention of getting employed; as “retired” if they had obtained retirement from service; as “unemployed” if they were not in any meaningful employment without receiving wages.

#### Statistical analysis

R-software was used to compile and analyze the data R, (2013). The normality of distribution of age and education distribution was verified using the Kolmogorov-Smirnov test. Propensity score matching was done using the “MatchIt” function in R (Ho, Imai, King, & Stuart, 2011). A propensity score was estimated for each subject in the sample based on a binary logistic regression model, with the “group status” (unaffected sibling in ADBS vs NMHS control) as the dependent variable, age and gender as the predictors. The Mahanalobis distances were then calculated between each subject in the unaffected sibling and the NMHS control group. Then, the “nearest neighbor” matching method was performed, to assign one NMHS control subject as a match for each unaffected sibling in the ADBS (1:1 ratio).

As more than one affected/unaffected individual participated from a given family, and siblings were at different stages of life-cycle, we analyzed affected and unaffected siblings as two independent groups. ANOVA and chi-square tests were applied to compare the variables across the three groups (affected / unaffected / NMHS controls).

## RESULTS

The study consisted of 259 affected siblings and 229 unaffected siblings from 202 multiplex families and 229 healthy controls. The age, gender and place of residence were matched across the groups. The affected and unaffected siblings had poorer educational attainment compared to healthy controls (F=8.9, p<0.001). Also, affected siblings were more likely to be single (χ2=40.97, p<0.001) and unemployed (χ2=39.776, p<0.001) in comparison to both their unaffected siblings, and healthy controls. Further, the unaffected siblings too were more likely to be single (32.8%) compared to healthy controls (22.7%). Affected individuals were more often separated or divorced (5.4%) when compared with that of the unaffected siblings (2.2%), whereas healthy controls had no individuals being separated or divorced. However, the unaffected siblings were more likely to be employed (only 3.5% were unemployed), as compared to that of affected siblings (18.5% were unemployed) and even healthy controls (8.7%). Female participants had fewer years of education, and were largely married, and were homemakers across the three groups when compared to males. However, almost 40% of affected females were single or divorced, as were almost a fifth of unaffected female siblings (22%), as compared to 7% in the general population. More female siblings were employed, in comparison to the general population, as well as ill (females) subjects, suggesting that the economic burden had nudged them towards joining paid work. The above results are depicted in following Tables-1, 2 and 3.

**Table 1:** Comparison of socio-demographic details between Affected Siblings, Unaffected Siblings and NMHS Healthy controls.

| Variables | | Affected<br>(N=259) | Unaffected<br>(N=229) | NMHS data<br>set (N=229) | $X^2/F^\#$ | P value | Post-hoc significant results |
| --- | --- | --- | --- | --- | --- | --- | --- |
| Age (in years) |  | 35.17<br>(10.25) | 34.64<br>(10.62) | 34.82<br>(10.39) | 0.164<sup>#</sup> | 0.849 | - |
| Education (in years) |  | 9.88 (4.68) | 10.41 (4.87) | 11.58 (3.85) | 8.967<sup>#</sup> | <0.001 | Affected<NMHS (p<0.001),<br>Unaffected<NMHS (p=0.016) |
| Gender | Male | 158 (61%) | 128 (55.9%) | 131 (57.2%) | 1.429 | 0.489 |  |
|  | Female | 101 (39%) | 101 (44.1%) | 98 (42.8%) |  |  |  |
| Marital<br>Status | Married | 134 (51.7%) | 149 (65.1%) | 177 (77.3%) | 40.978 | <0.001 | Affected vs. Unaffected (p=0.006)<br>Unaffected vs. NMHS (p=0.003)<br>Affected vs. NMHS (p<0.001) |
|  | Single | 111 (42.9%) | 75 (32.8%) | 52 (22.7%) |  |  |  |
|  | Separated/<br>Divorced | 14 (5.4%) | 5 (2.2%) | 0 (0%) |  |  |  |
| Place of<br>residence | Urban | 137 (53.1%) | 136 (59.6%) | 126 (55%) | 2.188 | 0.335 |  |
|  | Rural | 122 (46.9%) | 93 (40.4%) | 103 (45%) |  |  |  |
| Occupation | Employed | 142 (54.8%) | 156 (68.1%) | 130 (56.8%) | 39.776 | <0.001 | Affected vs. Unaffected (p<0.001)<br>Unaffected vs. NMHS (p=0.015)<br>Affected vs. NMHS (p=0.013) |
|  | Unemployed | 48 (18.5%) | 8 (3.5%) | 20 (8.7%) |  |  |  |
|  | Homemaker | 52 (20.1%) | 43 (18.8%) | 59 (25.8%) |  |  |  |
|  | Retired | 0 (0%) | 0 (0%) | 2 (0.9%) |  |  |  |
|  | Student | 17(6.6%) | 22 (9.6%) | 18 (7.9%) |  |  |  |

**Table 2:** Gender-wise comparisons within males only, across the three groups.

| | | Affected (n=158) | | Unaffected (n=128) | | NMHS (n=131) | | $\chi^2/F^\#$ | P value | Post-hoc significant results |
| --- | --- | --- | --- | --- | --- | --- | --- | --- | --- | --- |
|  |  | n/Mean | %/SD | n/Mean | %/SD | n/Mean | %/SD |  |  |  |
| Age (in years) |  | 33.64 | 9.76 | 33.95 | 10.01 | 33.82 | 9.93 | 0.035<sup>#</sup> | 0.966 | - |
| Education (in years) |  | 10.16 | 4.46 | 11.36 | 4.33 | 12.31 | 3.36 | 9.951<sup>#</sup> | <0.001 | Affected<NMHS (p<0.001)<br>Unaffected<NMHS (p=0.043) |
| Marital Status | Married | 75 | 47.5% | 71 | 55.5% | 86 | 65.6% | 12.33 | 0.015 | Only Affected vs. NMHS (p=0.002) |
|  | Single | 78 | 49.4% | 55 | 43% | 45 | 34.4% |  |  |  |
|  | Divorced or separated | 5 | 3.2% | 2 | 1.6% | 0 | 0% |  |  |  |
| Place of residence | Urban | 92 | 58.6% | 76 | 59.4% | 69 | 52.7% | 1.459 | 0.482 | - |
|  | Rural | 66 | 41.4% | 52 | 40.6% | 62 | 47.3% |  |  | - |
| Occupation | Employed | 119 | 75.3% | 111 | 86.7% | 101 | 77.1% | 18.68 | 0.017 | Only Affected vs. Unaffected (p=0.001) |
|  | Unemployed | 28 | 17.7% | 5 | 3.9% | 15 | 11.5% |  |  |  |
|  | Retired | 0 | 0% | 0 | 0% | 1 | 0.8% |  |  |  |
|  | Homemaker | 2 | 1.3% | 0 | 0% | 1 | 0.8% |  |  |  |
|  | Student | 9 | 5.7% | 12 | 9.4% | 13 | 9.9% |  |  |  |

**Table 3:** Gender-wise comparisons within females only, across the three groups.

|  |  | Affected (n=101) |  | Unaffected (n=101) |  | NMHS (n=98) |  | X<sup>2</sup>/F<sup>#</sup> | P value | Post-hoc significant results |
| --- | --- | --- | --- | --- | --- | --- | --- | --- | --- | --- |
|  |  | n/Mean | %/SD | n/Mean | %/SD | n/Mean | %/SD |  |  |  |
| Age (in years) |  | 37.56 | 10.60 | 35.52 | 11.33 | 36.15 | 10.90 | 0.919<sup>#</sup> | 0.4 | - |
| Education (in years) |  | 9.45 | 5.00 | 9.21 | 5.26 | 10.61 | 4.25 | 2.367<sup>#</sup> | 0.095 | - |
| Marital Status | Married | 59 | 58.4% | 78 | 77.2% | 91 | 92.9% | 34.283 | <0.001 | All three significantly different<br>Affected vs. Unaffected (p=0.012)<br>Unaffected vs. NMHS (p=0.006)<br>Affected vs. NMHS (p<0.001) |
|  | Single | 33 | 32.7% | 20 | 19.8% | 7 | 7.1% |  |  |  |
|  | Divorced or separated | 9 | 8.9% | 3 | 3% | 0 | 0% |  |  |  |
| Place of residence | Urban | 45 | 44.6% | 60 | 60% | 57 | 58.2% | 5.760 | 0.056 | - |
|  | Rural | 56 | 55.4% | 41 | 40% | 41 | 41.8% |  |  | - |
| Occupation | Employed | 23 | 22.8% | 45 | 44.6% | 29 | 29.6% | 32.185 | <0.001 | Affected vs. Unaffected (p<0.001)<br>Affected vs. NMHS (p=0.018) |
|  | Unemployed | 20 | 19.8% | 3 | 3% | 5 | 5.1% |  |  |  |
|  | Retired | 0 | 0% | 0 | 0% | 1 | 1% |  |  |  |
|  | Homemaker | 50 | 49.5% | 43 | 42.6% | 58 | 59.2% |  |  |  |
|  | Student | 8 | 7.9% | 10 | 9.9% | 5 | 5.1% |  |  |  |

## DISCUSSION

Individuals with severe mental illness tended to have poorer educational attainment, and more likely to be single and unemployed. Their siblings too show deficiencies in these parameters, except for employment.

The educational attainment and rates of marriage were better in unaffected siblings compared to their affected siblings; but it was still substantially poorer when compared to healthy controls. However, the unaffected siblings were likely to be employed, even when compared to healthy controls. This suggests that even though the unaffected siblings had poorer educational attainment, they had to bear the economic burden of the family by having to be in employment.

Female participants, in general, had spent fewer years in education. Although females tended to be married across three groups, those affected were more likely to have been divorced or separated. A substantially higher proportion of unaffected female siblings were employed compared to affected counterparts and healthy controls, suggesting that they had to take on the role of earning person and contributed financially to the family.

First-degree relatives, more so siblings of patients with psychiatric illness, are key in identifying the presumed genetic risk by identifying the vulnerability markers, even in the absence of any psychiatric symptoms (Jacob, 2016). Studies have evaluated the caregiver burden in family members with one affected member with psychiatric illness. The burden of illness in the family was thus apparent, even in unaffected siblings of those with severe mental illnesses. The impact could be driven by the biological and sociocultural factors influencing the functional abilities and markers of functioning (Subho Chakrabarti, 2016; S. Chakrabarti & Kulhara, 1999; Fujino & Okamura, 2009).

Academic underperformance can precede the onset of illness in schizophrenia and, bipolar disorder (Dickson & Hedges, 2020; Dickson, Laurens, Cullen, & Hodgins, 2012). Academic underachievement in unaffected siblings could reflect cognitive impairment as an endophenotype (Tempelaar, Termorshuizen, MacCabe, Boks, & Kahn, 2017), perhaps resulting from underlying genetic vulnerability (Toulopoulou et al., 2007) and also influenced by neurodevelopmental and environmental factors (Bora, 2015; Sirin, 2005).

In an earlier twin study, it was found that twins with a history of co-twin affected with bipolar disorder had poor educational and employment status compared to twins without a history of co-twin with an affected status (Christensen, Kyvik, & Kessing, 2007). A recent Danish nationwide population-based longitudinal study compared bipolar disorder patients and their siblings. They found that patients with bipolar disorder and their siblings had poor socio-occupational functioning and a decreased ability to improve their occupational and economic status on longitudinal follow-up compared to controls (Sletved, Ziersen, Andersen, Vinberg, & Kessing, 2021). This is, however, in contrast to our current study in which we found that unaffected siblings had better employment rates than the healthy controls from the NMHS data. Unemployment rates in affected siblings were higher than that in healthy controls, which was also found similar in a previous research study (Ramasubramanian, Mohandoss, & Namasivayam, 2016). Whether this reflects a greater proportion of ‘subsistence’ driven employment needs to be assessed. The Census of India (2011) also provides the same insight of lower employment status among mentally ill, perhaps due to residual deficits and the associated stigma. Deficits in education, cognition and social skills would contribute to the lower employment achievements in individuals with severe mental illness. The increased complexity of jobs in the post-industrialization era makes it even more difficult to maintain employment. A two-way relationship also exists; it could be that the poor socio-occupational determinants in affected individuals are a result of mental illness, or vice-versa (i.e., poor social-occupational determinants influencing mental illness in the affected individuals) (Allen, Balfour, Bell, & Marmot, 2014).

Marriage is another complex functional indicator, influenced by social factors. Affected and affected individuals were more likely to have remained unmarried or experienced marital difficulties. Illness characteristics such as early onset, the severity of the symptoms and greater disability results in dysfunctional marital and interpersonal relationships (Chopra, Bhaskaran, & Varma, 1970) (Hakulinen et al., 2019; Motovsky & Pecenak, 2013; Nisha, Sathesh, Punnoose, & Varghese, 2015). The sexual and reproductive functioning are impacted by severe mental illness, and contribute to marriage related issues. Females tend to get married at a young age and remain as homemakers when compared to males (Lowe, Joof, & Rojas, 2019) (Koujalgi & Patil, 2013), whereas males tend to remain single. However, the divorce rates were more in affected females (9%) than unaffected females (3%) and males across the three groups in the current study. Females with mental illness were thus much more likely to be ‘dumped’, thus affirming the stigma and gender bias in society.

A substantial proportion of unaffected female siblings were also employed compared to affected counterparts and healthy controls, suggesting that they had taken the role of earning person and contribute financially to the family. The proportion of women in employment in India is around 23% (https://data.worldbank.org/indicator/SL.TLF.TOTL.FE.ZS?locations=IN), and we see that a slightly higher proportion of healthy women (29%) were in paid employment when compared to those ill (22%); but almost half the female unaffected relatives were in paid employment (44%). The employment status of the men (80% of males between 18-64 years are expected to be employed [https://data.worldbank.org/indicator/SL.TLF.ACTI.MA.ZS?locations=IN]) is in the expected range and shows a similar trend of enhanced employment rates in unaffected siblings, although the magnitude of difference was clearly lower when compared with the analysis among female participants.

A greater proportion of females in the study reported their occupational status as homemakers. Societal norms regarding gender roles may have compromised some data. Unmarried women staying at home were often identified as being unemployed (though they may have been taking on domestic responsibilities), while married ones were designated as homemakers. In contrast, a male who is unmarried and under any form of employment, would be classified as ‘employed’. This could possibly explain the gender difference in our findings in employment. The other limitation is that the occupational status obtained in our study may not actually reflect the functional status of the individuals. This should be kept in mind while interpreting the results.

## CONCLUSION

Our study findings reveal the socio-demographic characteristics of affected and unaffected siblings from multiple affected families with major psychiatric illness. Affected and unaffected siblings had lower educational attainment and rates of marriage when compared to healthy controls. The unaffected siblings were more likely to be in employment, even when compared to healthy controls, and this finding was more prominent in females. Whether the poorer educational attainment and marital functioning in unaffected siblings is a biological marker of shared endophenotype or the effect of social burden (and stigma) of having an affected family member requires further evaluation. The support needs of these ‘high-risk’ families, whether in terms of disability support, housing or skilling, may need extra attention.

## Data Availability

All data produced in the present work are contained in the manuscript

## Acknowledgements

We thank all investigators of the National Mental Health Survey of India, for sharing their dataset for this analysis

## Notes

**Declaration of Interests:** All authors declare that they have no conflicts of interests to declare with respect to the current study.

### Competing Interest Statement

The authors have declared no competing interest.

### Funding Statement

This research is funded by the Accelerator program for Discovery in Brain disorders using Stem cells (ADBS) (jointly funded by the Department of Biotechnology, Government of India, and the Pratiksha trust; Grant BT/PR17316/MED/31/326/2015). BV is funded by the Intermediate (Clinical and Public Health) Fellowship (IA/CPHI/20/1/505266) of the DBT/Wellcome Trust India Alliance.

### Author Declarations

The studies were approved by the Institutional ethics committee (NIMHANS Ethics committee). NIMHANS Ethics committee gave ethical approval for this work.

